# Comparative effectiveness of COVID-19 vaccination against death and severe disease in an ongoing nationwide mass vaccination campaign

**DOI:** 10.1101/2022.01.28.22270009

**Authors:** Theodore Lytras, Flora Kontopidou, Angeliki Lambrou, Sotirios Tsiodras

## Abstract

**Background:** As national COVID-19 mass vaccination campaigns are rolled out, it is important to demonstrate and measure their public health benefit. We aimed to estimate COVID-19 Vaccine Effectiveness (VE) against severe disease and death in the Greek population, for all vaccines in use.

**Methods:** Nationwide active surveillance and vaccination registry data during January-December 2021 were used to estimate VE via quasi-Poisson regression, as one minus the Incidence Rate Ratio, adjusted for age and calendar time. Interaction terms were included to assess VE by age group, against the “delta” SARS-CoV-2 variant and waning of VE over time.

**Results:** Two doses of BNT162b2, mRNA-1273 or ChAdOx1 nCov-19 vaccines offered very high (>90%) VE against both intubation and death across all age groups, similar against both “delta” and previous variants, with one-dose Ad26.COV2.S slightly lower. There was some waning over time but VE remained >80% at six months, and three doses increased VE again to near 100%. Vaccination prevented an estimated 19,691 COVID-19 deaths (95% CI: 18,890-20,788) over the study period.

**Conclusions:** All approved vaccines were very highly effective in preventing COVID-19 severe disease and death. Every effort should be made to vaccinate the population with at least two doses, in order to reduce the mortality and morbidity impact of the pandemic.

## Introduction

The efficacy of currently authorized Coronavirus disease 2019 (COVID-19) vaccines in preventing symptomatic infection has been well documented in phase 3 clinical trials [1–4] and further demonstrated in multiple observational studies [5–8]. However, as the Severe Acute Respiratory Syndrome Coronavirus 2 (SARS-CoV-2) transitions towards endemicity [9], preventing severe COVID-19 through vaccination becomes a strategic priority to reduce overall disease burden and mortality and safeguard healthcare services [10]. It is therefore vitally important to monitor COVID-19 Vaccine Effectiveness (VE) against severe disease and death in a real life setting, to quantify the Public Health benefit of vaccination, generate confidence and promote uptake. Long-term observational VE studies can further provide much-needed answers about the duration of protection and its extent against different SARS-CoV-2 variants.

Since December 2020 Greece has rolled out a centralized nationwide mass vaccination campaign, offering four COVID-19 vaccines to its entire resident population free-of-charge; two mRNA vaccines, BNT162b2 (Pfizer-BioNTech) and mRNA-1273 (Moderna), and two adenoviral vector vaccines, ChAdOx1 nCoV-19 (AstraZeneca) and Ad26.COV2.S (Janssen). This offers the opportunity to study the comparative effectiveness of these vaccines in the same population and setting. Roll-out was staggered by age group between January and July 2021, with healthcare workers and the elderly getting vaccinated first (Supplementary Table 1). Since September 2021 a third mRNA vaccine dose was offered to all persons who completed two-dose vaccination six months prior.

**Table 1:** Crude and age-adjusted rates of COVID-19 intubation and death by vaccination group, Greece, 1 January to 9 December 2021.

| Vaccination group* | Person-years of follow-up | Median follow-up, days (IQR) | Median age, years (IQR)** | Intubations |  |  | Deaths |  |  |
| --- | --- | --- | --- | --- | --- | --- | --- | --- | --- |
|  |  |  |  | Number of intubations | Rate per 100k population | Age-adjusted rate per 100k population | Number of deaths | Rate per 100k population | Age-adjusted rate per 100k population |
| Unvaccinated | 5138482 | N/A | 45 (30-60) | 7984 | 155.38 | 92.82 | 12172 | 236.88 | 141.52 |
| 3-dose (any vaccine) | 106773 | 15 (6-30) | 75 (61-81) | 24 | 22.48 | 0.28 | 52 | 48.70 | 0.60 |
| 2-dose (any vaccine) | 2638990 | 167 (133-191) | 58 (43-72) | 684 | 25.92 | 7.95 | 1806 | 68.44 | 21.00 |
| 1-dose (any vaccine) | 720464 | 21 (20-27) | 50 (35-63) | 408 | 56.63 | 4.74 | 725 | 100.63 | 8.43 |
| 2-dose BNT162b2 | 2037824 | 168 (131-194) | 57 (43-74) | 548 | 26.89 | 6.37 | 1629 | 79.94 | 18.94 |
| 2-dose ChAdOx1 nCov-19 | 341120 | 166 (150-185) | 61 (43-64) | 101 | 29.61 | 1.17 | 133 | 38.99 | 1.55 |
| 1-dose BNT162b2 | 331326 | 20 (20-21) | 52 (36-68) | 272 | 82.09 | 3.16 | 499 | 150.61 | 5.80 |
| 2-dose mRNA-1273 | 251492 | 175 (149-194) | 55 (44-70) | 35 | 13.92 | 0.41 | 42 | 16.70 | 0.49 |
| 1-dose Ad26.COV2.S | 184317 | 148 (90-176) | 39 (26-49) | 41 | 22.24 | 0.48 | 110 | 59.68 | 1.28 |
| 1-dose ChAdOx1 nCoV-19 | 157366 | 76 (61-79) | 61 (43-64) | 66 | 41.94 | 0.77 | 87 | 55.29 | 1.01 |
| 3-dose BNT162b2 | 91089 | 18 (7-35) | 75 (60-82) | 24 | 26.35 | 0.28 | 51 | 55.99 | 0.59 |
| 1-dose mRNA-1273 | 47455 | 27 (27-28) | 52 (40-67) | 29 | 61.11 | 0.34 | 29 | 61.11 | 0.34 |

\* Only combinations with at least 10 events (deaths or intubations) and 10,000 person-years of associated follow-up.
\*\* Distribution weighted by follow-up time.

The objective of this observational study was to estimate COVID-19 VE against severe disease and death for all available vaccine/dose combinations, stratified by age group, in the entire Greek population. Secondary objectives were to detect waning of VE over time, and to estimate VE against the “delta” SARS-CoV-2 variant (vs previously circulating variants).

## Methods

### Study population and data collection

In Greece the National Public Health Organization (EODY) is responsible for COVID-19 surveillance, with particular emphasis on active case finding and follow-up of all severe cases nationwide. A severe COVID-19 case is defined as a laboratory-confirmed case (using PCR or antigen testing) that has been intubated and/or hospitalized in intensive care or has died (regardless of setting). As the availability of intensive care beds varied during the pandemic, we focused on intubation and death as the two most unbiased and completely ascertained outcomes. COVID-19 deaths were defined under World Health Organization guidelines as those with laboratory confirmation and clinically compatible illness without complete recovery preceding death, and with no time cut-off following laboratory confirmation [11].

For the study period between 11 January 2020 and 8 December 2021 we obtained anonymized surveillance data for all COVID-19 intubations and deaths in Greece in persons aged 15 years and older (9.2 million population), including their vaccination status at the time of laboratory confirmation as reported to EODY by the National Vaccine Registry (NVR). The NVR is operated by the Hellenic Ministry of Digital Governance and registers all COVID-19 vaccinations performed in Greece, without exception. From the NVR we additionally obtained aggregate counts of persons vaccinated with every vaccine/dose combination, for every age and every day during the study period, and further stratified by time since vaccination; this allowed accurate measurement of person-time spent in each vaccination “state”, and thus calculation of crude and age-adjusted rates of COVID-19 intubations and deaths for each vaccination group, based on the dates of laboratory confirmation (rather than dates of intubation or death).

No funding was received for this study. The study was approved by the EODY board; as only anonymized data were used from which no person can be identified, no separate ethical approval was required. The board had no role in study design, analysis, interpretation of data and decision to publish.

### Statistical analysis

VE against COVID-19 intubation and against COVID-19 death were estimated using quasi-Poisson regression as one minus the Incidence Rate Ratio (IRR), adjusted for age (in 5-year groups) and calendar time (calendar week as categorical variable). Two models were fitted: model A, grouping all vaccines together and comparing 1-dose, 2-dose and 3-dose vaccination to the unvaccinated, and model B, assessing separately each vaccine/dose combination among those that had at least 10 events and 10,000 person-years of associated follow-up. Both models included additional interaction terms: (a) between vaccine and age (15-59, 60-79 and 80+ years) to detect any diminished effectiveness among the elderly; (b) between vaccine and time since one month after the last received dose, in order to assess waning of VE compared to the first month after vaccination; and (c) between vaccine and prevalence of the “delta” SARS-CoV-2 variant. Data on the latter were provided by the National SARS-CoV-2 Genomic Surveillance Network, coordinated by EODY; before ISO week 25/2021 “delta” was absent and after week 30/2021 fully dominant, while for the intermediate period (weeks 25-30/2021) the proportion of “delta” among all randomly selected and genotyped samples was used (Supplementary Table 2). In case of severe collinearity (Variance Inflation Factor over 5) or sparse data (<5 tabulated events) the respective interaction terms were dropped from model B. Also the interaction term between vaccine and time since last dose was dropped for 3-dose vaccination as follow-up time was too short (Supplementary Table 3). Stratified VE estimates were calculated as linear combinations of the respective model coefficients. We additionally used the fitted models to estimate the number of intubations and deaths prevented by vaccination during the study period, as the predicted number of outcome events if everyone was unvaccinated, minus observed events [12]. All statistical analyses were performed with the R statistical environment, version 4.1.2 [13].

## Results

During the study period, a total of 14,676,605 vaccine doses were administered in Greece (11,427,784 BNT162b2, 1,161,905 mRNA-1273, 1,505,334 ChAdOx1 nCoV-19 and 581,582 Ad26.COV2.S) and a total of 9100 COVID-19 intubations and 14755 COVID-19 deaths occurred. Event rates and follow-up time per vaccination group are detailed in Table 1; even though a substantial number of vaccinated persons died of COVID-19, crude and age-adjusted rates were several times lower compared to the unvaccinated group. Vaccinated individuals, especially those that received 3 doses, were also older on average than the unvaccinated (Table 1). Weekly rates of COVID-19 intubation and death varied greatly over time, reflecting SARS-CoV-2 community prevalence and spread (Figure 1).

**Figure 1:**
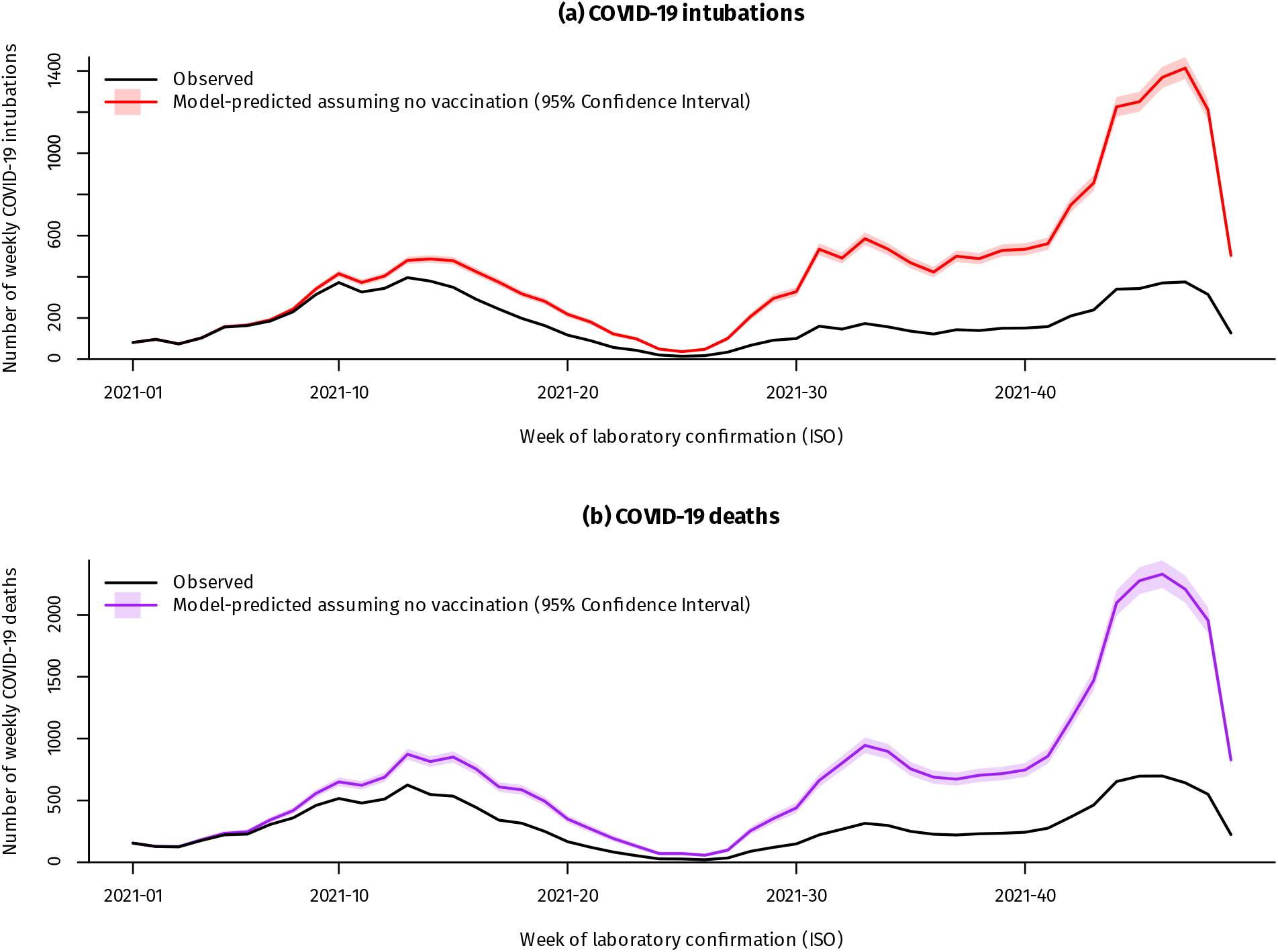
Weekly COVID-19 deaths and intubations in Greece, January-December 2021, observed vs model-predicted assuming no vaccination. ***Footnote for Figure 1:*** The latest weeks in the series are subject to reporting delay.

The most relevant adjusted VE estimates from models A and B are presented in Figures 2 and 3 respectively (full results are detailed in the online supplement). Two COVID-19 vaccine doses offered more than 90% protection against death and intubation across all age groups, with marginally lower protection in persons 80 years or older. VE against death was similar for both “delta” and previously circulating variants, and even slightly higher against intubation from the “delta” variant (p<0.001). However, 2-dose VE waned slightly over time while still being over 80% at 6 months, and a third vaccine dose restored it close to 100% even in the oldest age group. Effectiveness for one vaccine dose was suboptimal but still sufficient to halve the rate of both death and intubation compared to the unvaccinated (Figure 2). Both the BNT162b2, mRNA-1273 and ChAdOx1 nCoV-19 vaccines showed similar and very high VE (>90%) in all age groups, with a slight waning over time that was marginally better with mRNA-1273 particularly among younger adults (Figure 3). In contrast, one dose of Ad26.COV2.S offered significantly less protection against intubation or death during the first month after vaccination, similar to that of one dose of BNT162b2; over time, however, VE for Ad26.COV2.S increased and at 6 months was comparable to that of the other 2-dose vaccines (Figure 3).

**Figure 2:**
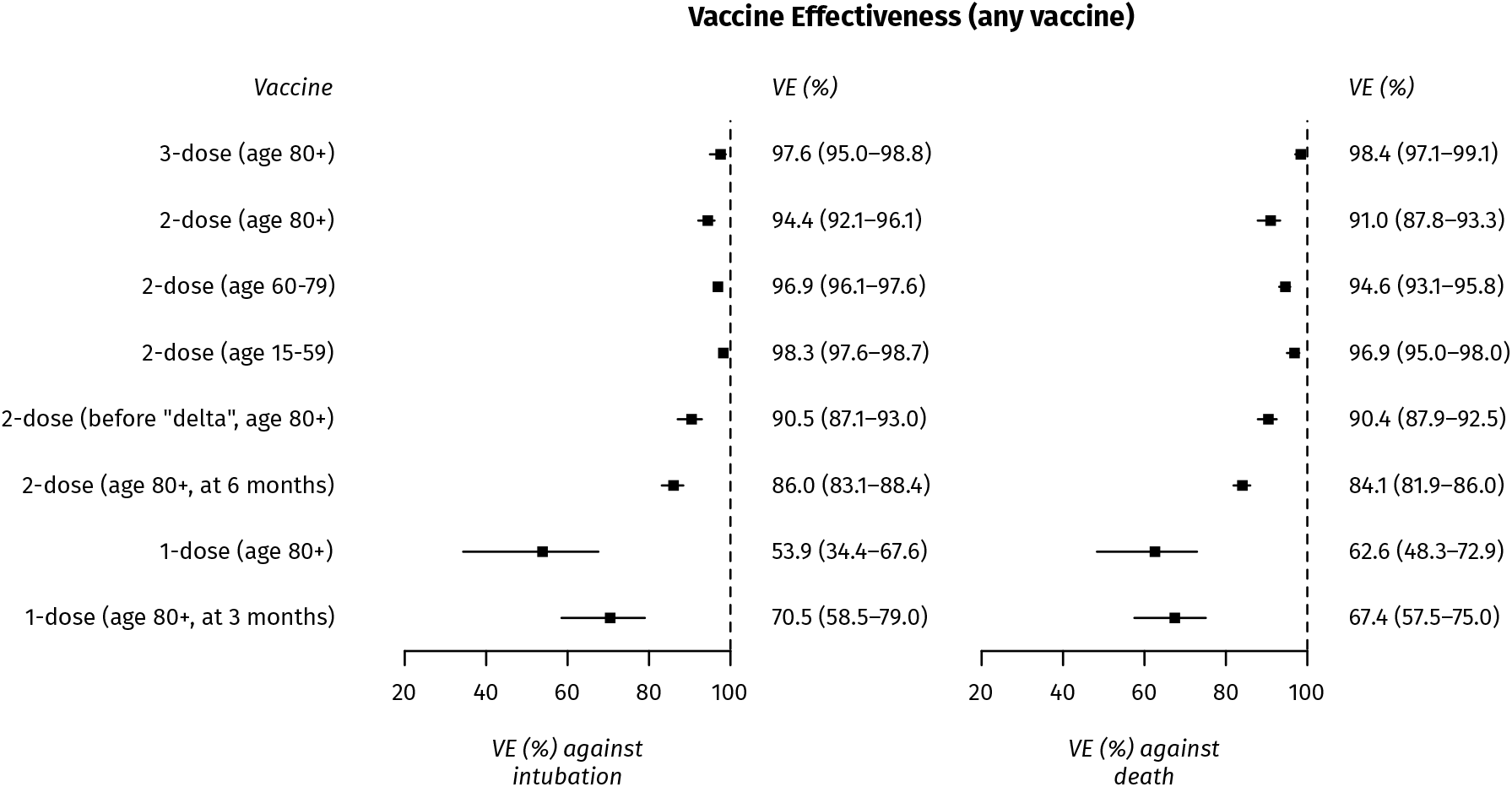
Effectiveness of 1-, 2- and 3-dose vaccination against COVID-19 intubation and death, Greece, January-December 2021 (model A)

**Figure 3:**
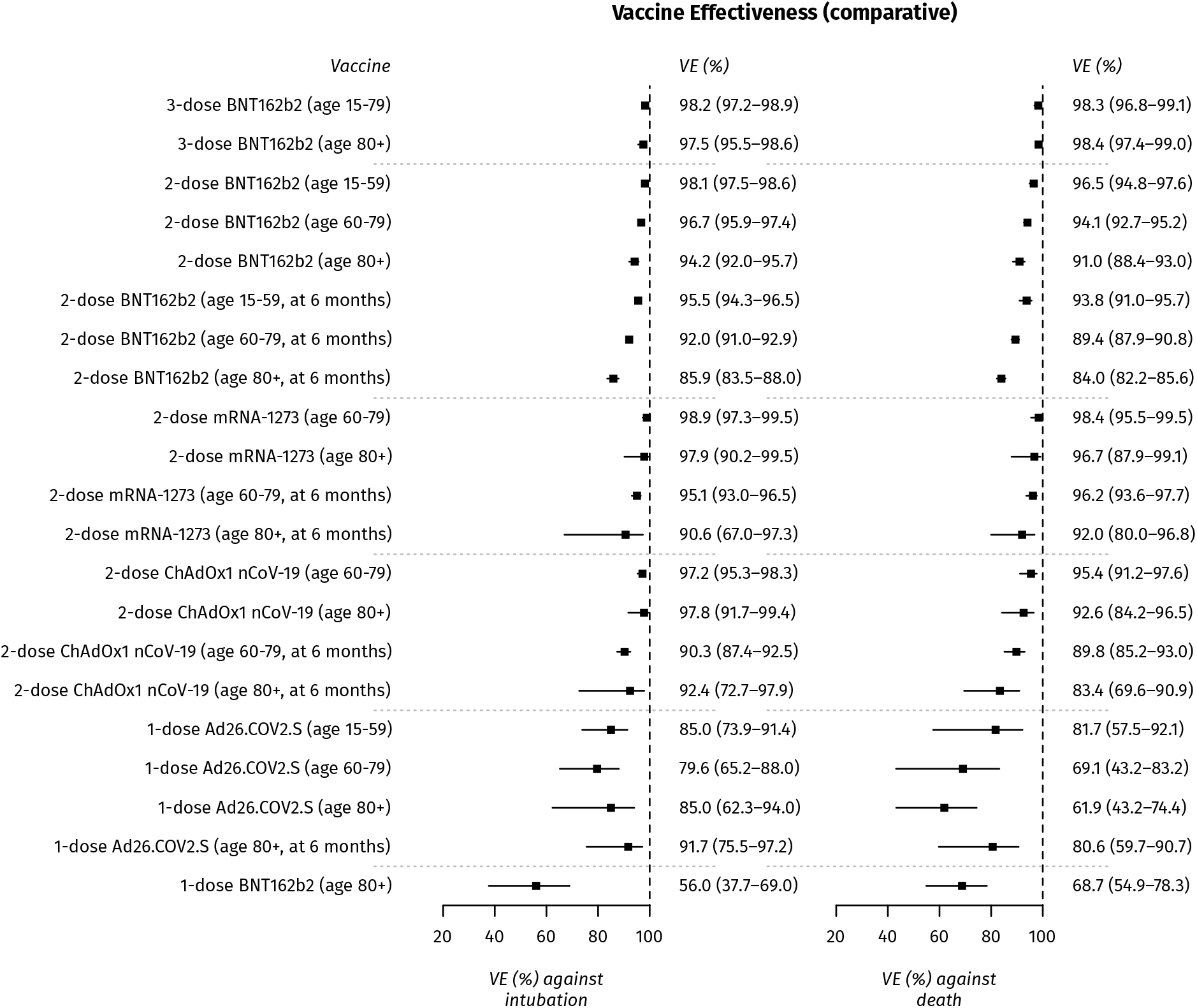
Comparative effectiveness of BNT162b2, mRNA-1273, ChAdOx1 nCoV-19 and Ad26.COV2.S vaccines against COVID-19 intubation and death, Greece, January-December 2021 (model B)

Given the above associations from model B, if vaccination was not available the pandemic would have had a much more severe impact, especially during the large “delta” wave of winter 2021 (Figure 1); over the entire study period, vaccination prevented an estimated 19,691 COVID-19 deaths (95% CI: 18,890-20,788) and 6,674 COVID-19 intubations (95% CI: 6,251-7,241). Finally, there was a very steep age gradient in the rate of COVID-19 death, with more than a thousand-fold difference between the younger and older age groups; a similar gradient was observed for intubations but the rate was lower in people older than 80 years, suggesting clinicians may avoid invasive mechanical ventilation in severely ill elderly patients [14] (Figure 4).

**Figure 4:**
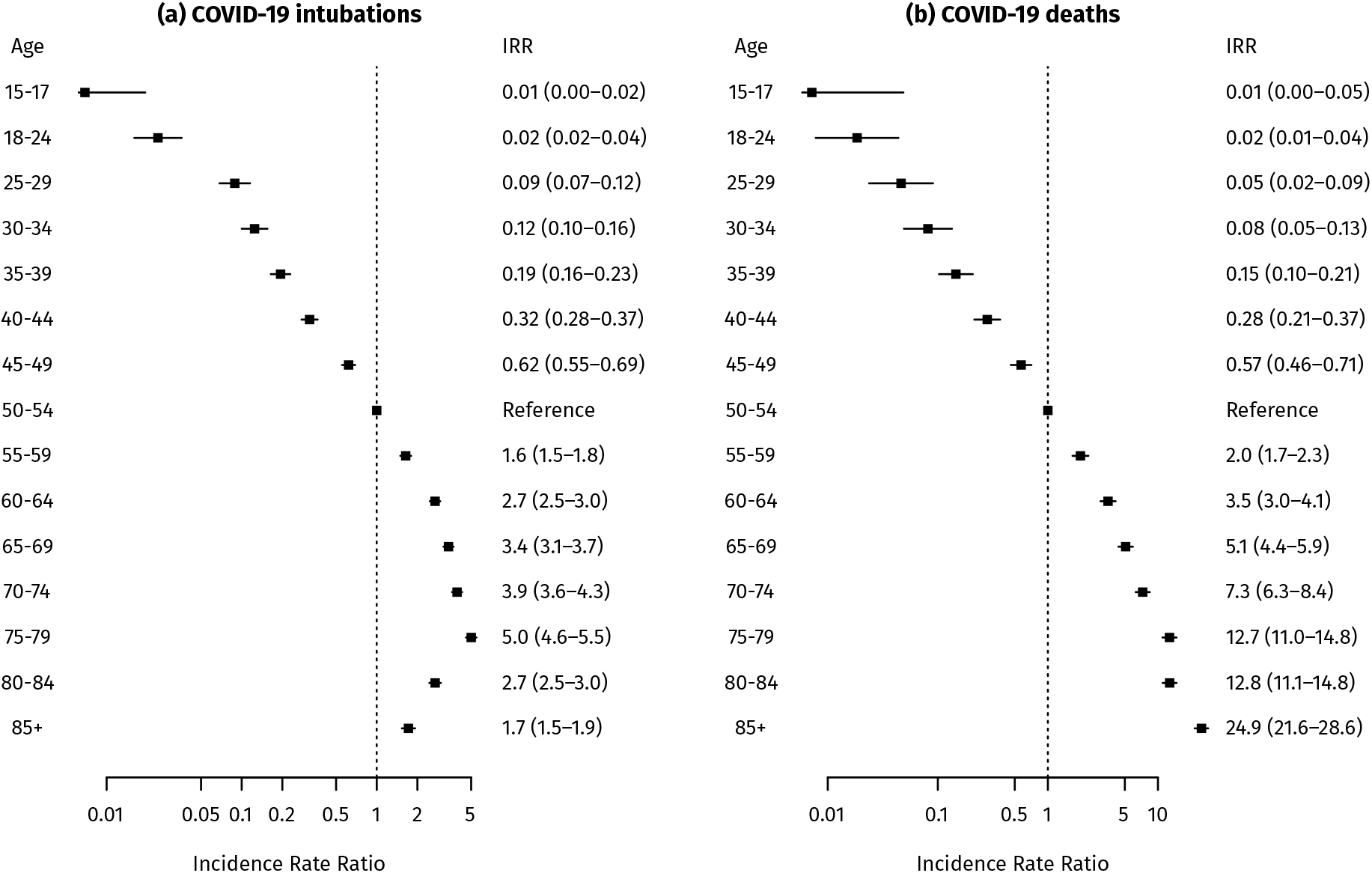
Rates of COVID-19 intubation and death by age group, Greece, January-December 2021 (model B)

## Discussion

Our study provides important real-world validation for the remarkable effectiveness of COVID-19 vaccination against severe disease or death and quantifies the public health benefit from vaccination. It demonstrates that VE is similar and very high (>90%) across all age groups, young and old, despite the absolute benefit being higher for older people due to their much higher susceptibility. Furthermore, effectiveness was maintained (and for severe disease, even increased) against the “delta” variant of SARS-CoV-2 compared to “alpha” and older variants, as seen in other populations [15–17]. The findings confirm that vaccination is by far the most important public health tool to blunt the mortality and morbidity impact of the COVID-19 pandemic, prevent overloading of healthcare services and save lives [18].

With the large size and long follow-up of our study population, we were able to demonstrate a small, yet statistically significant, waning of effectiveness against severe disease and death. Even in the oldest age group though, VE six months after two doses of mRNA or ChAdOx1 nCoV-19 vaccine remained higher than 80%, a very substantial level of protection, which is further amplified by a third dose to levels even higher than the initial 2-dose vaccination. Our findings are very similar to a recent study from England using the test-negative design [19]. This has implications for vaccine rollout; as Greece and other countries completed 2-dose vaccination of most middle-aged and older adults before summer 2021, it was possible to optimize their protection by offering a third dose six months later, just ahead of the anticipated winter pandemic wave. On the other hand, the difference in VE between 2-dose and 3-dose vaccination is much smaller than between 1-dose and 2-dose, or 1-dose and being unvaccinated; therefore for public health authorities, closing the existing COVID-19 vaccination gaps is an even more urgent priority than offering third doses to the population [20, 21]. At the same time, the benefit from a third dose needs to be balanced against the ethical and utilitarian need for global vaccine equity [22, 23]. However, an additional challenge is presented by the recent emergence of the “omicron” variant, given the substantial benefit of the third dose in preventing symptomatic infection as well as severe disease due to “omicron” [24].

The currently four vaccines approved and used in Greece show very similar effectiveness, with only marginal differences. Interestingly, VE for 1-dose Ad26.COV2.S is consistent with its post-vaccination antibody response, which is initially lower but more stable over time [25]. The similar VE for all vaccines is important, as there are no head-to-head randomized trials, and provides valuable reassurance to the public who might ask which vaccine is best. It also shifts considerations about specific vaccine recommendations away from effectiveness and primarily towards availability, logistics and safety profile issues.

Strengths of our study include the large sample size, long follow-up and practically complete ascertainment of both vaccination and severe outcomes across the entire population of Greece, minimizing selection bias or bias due to healthseeking behaviour. Detailed information on age enabled tight adjustment for confounding; given the very large effect of age on the risk of severe COVID-19 disease and death, residual confounding by age is an important concern, and may lead to VE underestimation. Adjustments for calendar time and time since vaccination allowed accounting for waning effectiveness and the variable SARS-CoV-2 spread in the community, which was not always possible in other studies.

However, there are certain limitations as well. We did not examine laboratory-confirmed COVID-19 cases to estimate VE against infection, as these are highly dependent on healthseeking behaviour and testing patterns, and thus susceptible to bias. We did not have information on comorbidities or socioeconomic status; these are possible confounders since they are associated with both vaccination and risk of COVID-19 [26], although comorbidities are also largely associated with age, for which adjustment was made. Finally, we did not have information on previous SARS-CoV-2 infection; since infected persons were recommended to receive a single dose of COVID-19 vaccine, which creates strong “hybrid” immunity [27], this might have led to some overestimation of 1-dose VE in particular.

In conclusion, our study provides valuable documentation of the very high and durable effectiveness of COVID-19 vaccination in preventing severe disease and death in all age groups, both against the “delta” and older SARS-CoV-2 variants. The findings support the efforts to promote vaccination uptake, thereby reducing the mortality and morbidity impact of the COVID-19 pandemic. Conducting formal studies to monitor the effectiveness of COVID-19 vaccination is essential, as simple comparisons of counts or rates in surveillance data will underestimate effectiveness primarily due to confounding by age. Finally, as the “omicron” variant emerges, our study provides a blueprint for long-term monitoring of vaccine effectiveness during this new phase of the pandemic as well.

## Data Availability

The data produced in the present study are available upon reasonable request to the authors and after institutional approval.

## Author contributions

Original idea: TL, ST; Data analysis: TL; Data interpretation: TL, FK, AL, ST; First draft of the manuscript: TL; Revision of the manuscript for important intellectual content: TL, FK, AL, ST

## References

[1] Polack FP, Thomas SJ, Kitchin N, et al. Safety and Efficacy of the BNT162b2 mRNA Covid-19 Vaccine. N Engl J Med 2020; 383: 2603–2615.

[2] Baden LR, El Sahly HM, Essink B, et al. Efficacy and Safety of the mRNA-1273 SARS-CoV-2 Vaccine. N Engl J Med 2021; 384: 403–416.

[3] Falsey AR, Sobieszczyk ME, Hirsch I, et al. Phase 3 Safety and Efficacy of AZD1222 (ChAdOx1 nCoV-19) Covid-19 Vaccine. N Engl J Med. Epub ahead of print September 29, 2021. DOI: 10.1056/NEJMoa2105290.

[4] Sadoff J, Gray G, Vandebosch A, et al. Safety and Efficacy of Single-Dose Ad26.COV2.S Vaccine against Covid-19. N Engl J Med 2021; 384: 2187–2201.

[5] Bajema KL, Dahl RM, Prill MM, et al. Effectiveness of COVID-19 mRNA Vaccines Against COVID-19-Associated Hospitalization - Five Veterans Affairs Medical Centers, United States, February 1-August 6, 2021. MMWR Morb Mortal Wkly Rep 2021; 70: 1294–1299.

[6] Dagan N, Barda N, Kepten E, et al. BNT162b2 mRNA Covid-19 Vaccine in a Nationwide Mass Vaccination Setting. N Engl J Med 2021; 384: 1412–1423.

[7] Pilishvili T, Gierke R, Fleming-Dutra KE, et al. Effectiveness of mRNA Covid-19 Vaccine among U.S. Health Care Personnel. N Engl J Med. Epub ahead of print September 22, 2021. DOI: 10.1056/NEJMoa2106599.

[8] Thompson MG, Stenehjem E, Grannis S, et al. Effectiveness of Covid-19 Vaccines in Ambulatory and Inpatient Care Settings. New England Journal of Medicine 2021; 385: 1355–1371.

[9] Antia R, Halloran ME. Transition to endemicity: Understanding COVID-19. Immunity 2021; 54: 2172–2176.

[10] De Foo C, Grépin KA, Cook AR, et al. Navigating from SARS-CoV-2 elimination to endemicity in Australia, Hong Kong, New Zealand, and Singapore. Lancet 2021; 398: 1547–1551.

[11] World Health Organization. International guidelines for certification and classification (coding) of COVID-19 as cause of death., http://www.who.int/classifications/icd/Guidelines_Cause_of_Death_COVID-19.pdf (2020).

[12] Greenland S, Drescher K. Maximum likelihood estimation of the attributable fraction from logistic models. Biometrics 1993; 49: 865–872.

[13] R Core Team. R: A Language and Environment for Statistical Computing. Vienna, Austria: R Foundation for Statistical Computing, http://www.R-project.org/ (2019).

[14] Grasselli G, Zangrillo A, Zanella A, et al. Baseline Characteristics and Outcomes of 1591 Patients Infected With SARS-CoV-2 Admitted to ICUs of the Lombardy Region, Italy. JAMA 2020; 323: 1574–1581.

[15] Tang P, Hasan MR, Chemaitelly H, et al. BNT162b2 and mRNA-1273 COVID-19 vaccine effectiveness against the SARS-CoV-2 Delta variant in Qatar. Nat Med. Epub ahead of print November 2, 2021. DOI: 10.1038/s41591-021-01583-4.

[16] Bruxvoort KJ, Sy LS, Qian L, et al. Effectiveness of mRNA-1273 against delta, mu, and other emerging variants of SARS-CoV-2: test negative case-control study. BMJ 2021; 375: e068848.

[17] Barda N, Dagan N, Cohen C, et al. Effectiveness of a third dose of the BNT162b2 mRNA COVID-19 vaccine for preventing severe outcomes in Israel: an observational study. Lancet 2021; S0140-6736(21)02249–2.

[18] Meslé MM, Brown J, Mook P, et al. Estimated number of deaths directly averted in people 60 years and older as a result of COVID-19 vaccination in the WHO European Region, December 2020 to November 2021. Euro Surveill 2021; 26: 2101021.

[19] Andrews N, Tessier E, Stowe J, et al. Duration of Protection against Mild and Severe Disease by Covid-19 Vaccines. N Engl J Med. Epub ahead of print January 12, 2022. DOI: 10.1056/NEJMoa2115481.

[20] Finney Rutten LJ, Zhu X, Leppin AL, et al. Evidence-Based Strategies for Clinical Organizations to Address COVID-19 Vaccine Hesitancy. Mayo Clin Proc 2021; 96: 699–707.

[21] Razai MS, Chaudhry UAR, Doerholt K, et al. Covid-19 vaccination hesitancy. BMJ 2021; 373: n1138.

[22] Schaefer GO, Leland RJ, Emanuel EJ. Making Vaccines Available to Other Countries Before Offering Domestic Booster Vaccinations. JAMA 2021; 326: 903–904.

[23] The Lancet Infectious Diseases null. COVID-19 vaccine equity and booster doses. Lancet Infect Dis 2021; 21: 1193.

[24] Abu-Raddad LJ, Chemaitelly H, Ayoub HH, et al. Effectiveness of BNT162b2 and mRNA-1273 COVID-19 boosters against SARS-CoV-2 Omicron (B.1.1.529) infection in Qatar. 2022; 2022.01.18.22269452.

[25] Collier A-RY, Yu J, McMahan K, et al. Differential Kinetics of Immune Responses Elicited by Covid-19 Vaccines. N Engl J Med 2021; 385: 2010–2012.

[26] Williamson EJ, Walker AJ, Bhaskaran K, et al. Factors associated with COVID-19-related death using OpenSAFELY. Nature 2020; 584: 430–436.

[27] Goel RR, Painter MM, Apostolidis SA, et al. mRNA vaccines induce durable immune memory to SARS-CoV-2 and variants of concern. Science 2021; 374: abm0829.

