## Supplementary for "Comparative effectiveness of COVID-19 vaccination against death and severe disease in an ongoing nationwide mass vaccination campaign"

Supplementary Material for the article "**Comparative effectiveness of COVID-19 vaccination against death and severe disease in an ongoing nationwide mass vaccination campaign**"

**Supplementary Table 1:** Timeline of vaccine rollout in Greece

**Supplementary Table 2:** Proportion of the "delta" SARS-CoV-2 variant among randomly selected and genotyped SARS-CoV-2 samples, Greece, National SARS-CoV-2 Genomic Surveillance Network

**Supplementary Table 3:** Follow-up distribution per vaccination group (days)

**Supplementary Figure 1:** Effectiveness of 1-, 2- and 3-dose vaccination against COVID-19 death and intubation, Greece, January-December 2021 (full results – all vaccines grouped)

**Supplementary Figure 2:** Comparative effectiveness of BNT162b2, mRNA-1273, ChAdOx1 nCoV-19 and Ad26.COV2.S vaccines against COVID-19 death and intubation, Greece, January-December 2021 (full results – complete vaccinations)

**Supplementary Figure 3:** Comparative effectiveness of BNT162b2, mRNA-1273, ChAdOx1 nCoV-19 and Ad26.COV2.S vaccines against COVID-19 death and intubation, Greece, January-December 2021 (full results – incomplete vaccinations)

**Supplementary Table 1:** Timeline of vaccine rollout in Greece

| Date | Rollout |
|---|---|
| 20 December 2020 | Healthcare workers |
| 11 January 2021 | Ages 85+, all available vaccines |
| 22 January 2021 | Ages 80-84, all available vaccines |
| 10 February 2021 | Ages 60-64, ChAdOx1 nCoV-19 vaccine only |
| 12 February 2021 | Ages 75-79, all available vaccines |
| 26 March 2021 | Ages 70-74, all available vaccines |
| 2 April 2021 | Ages 65-69, all available vaccines |
| 10 April 2021 | Ages 60-64, all available vaccines |
| 21 April 2021 | Ages 50-54, all available vaccines |
| 24 April 2021 | Ages 55-59, all available vaccines |
| 27 April 2021 | Ages 30-39, ChAdOx1 nCoV-19 vaccine only |
| 29 April 2021 | Ages 40-44, all available vaccines |
| 1 May 2021 | Ages 45-49, all available vaccines |
| 26 May 2021 | Ages 35-39, all available vaccines |
| 29 May 2021 | Ages 30-34, all available vaccines<br>Ages 18+, Ad26.COV2.S vaccine only |
| 10 June 2021 | Ages 25-29, all available vaccines except ChAdOx1 nCoV-19 |
| 16 June 2021 | Ages 18-24, all available vaccines except ChAdOx1 nCoV-19 |
| 15 July 2021 | Ages 15-17, BNT162b2 vaccine only |
| 30 July 2021 | Ages 12-14, BNT162b2 vaccine only |
| 14 September 2021 | Third vaccine dose, for immunocompromised patients, BNT162b2 or mRNA-1273 vaccine only |
| 30 September 2021 | Third vaccine dose, healthcare workers and ages 60+, BNT162b2 or mRNA-1273 vaccine only |
| 10 October 2021 | Third vaccine dose, ages 50+, BNT162b2 or mRNA-1273 vaccine only |
| 5 November 2021 | Repeat dose for Ad26.COV2.S recipients, Ad26.COV2.S or BNT162b2 or mRNA-1273 vaccine |
| 20 November 2021 | Third vaccine dose, ages 18+, BNT162b2 or mRNA-1273 vaccine only |
| 10 December 2021 | Ages 5-11, pediatric BNT162b2 vaccine only |

The above timeline was determined by vaccine availability and the recommendations of the Greek National Commitee for Immunizations.

**Supplementary Table 2:** Proportion of the "delta" SARS-CoV-2 variant among randomly selected and genotyped SARS-CoV-2 samples, Greece, National SARS-CoV-2 Genomic Surveillance Network

| Week(s) | Samples with "Delta" variant | Samples with other variants | "Delta" proportion (%) |
|---|---|---|---|
| 20-24/2021 | 26 | 4,389 | 0.6 |
| 25/2021 | 20 | 249 | 8.0 |
| 26/2021 | 155 | 484 | 32.0 |
| 27/2021 | 741 | 1,341 | 55.3 |
| 28/2021 | 1,474 | 1,969 | 74.9 |
| 29/2021 | 1,435 | 1,630 | 88.0 |
| 30/2021 | 930 | 1,013 | 91.8 |
| 31-35/2021 | 3,795 | 3,861 | 98.3 |

Before week 25/2021, the most common circulating variants were B.1.1.7 ("Alpha") and B.1.1.318.

**Supplementary Table 3:** Follow-up distribution per vaccination group (days)

| | Quantile | | | | |
|---|---|---|---|---|---|
| **Vaccine group** | **2.5%** | **25%** | **50%** | **75%** | **97.5%** |
| 1-dose BNT162b2 | 0 | 6 | 12 | 18 | 121 |
| 1-dose mRNA-1273 | 0 | 7 | 15 | 23 | 120 |
| 1-dose ChAdOx1 nCoV-19 | 1 | 18 | 37 | 56 | 135 |
| 1-dose Ad26.COV2.S | 3 | 34 | 73 | 118 | 179 |
| 2-dose BNT162b2 | 4 | 41 | 85 | 134 | 222 |
| 2-dose mRNA-1273 | 4 | 43 | 86 | 132 | 197 |
| 2-dose ChAdOx1 nCoV-19 | 4 | 41 | 83 | 126 | 180 |
| 3-dose BNT162b2 | 0 | 6 | 15 | 30 | 59 |

**Supplementary Figure 1:** Effectiveness of 1-, 2- and 3-dose vaccination against COVID-19 death and intubation, Greece, January-December 2021 (full results, model A – all vaccines grouped)

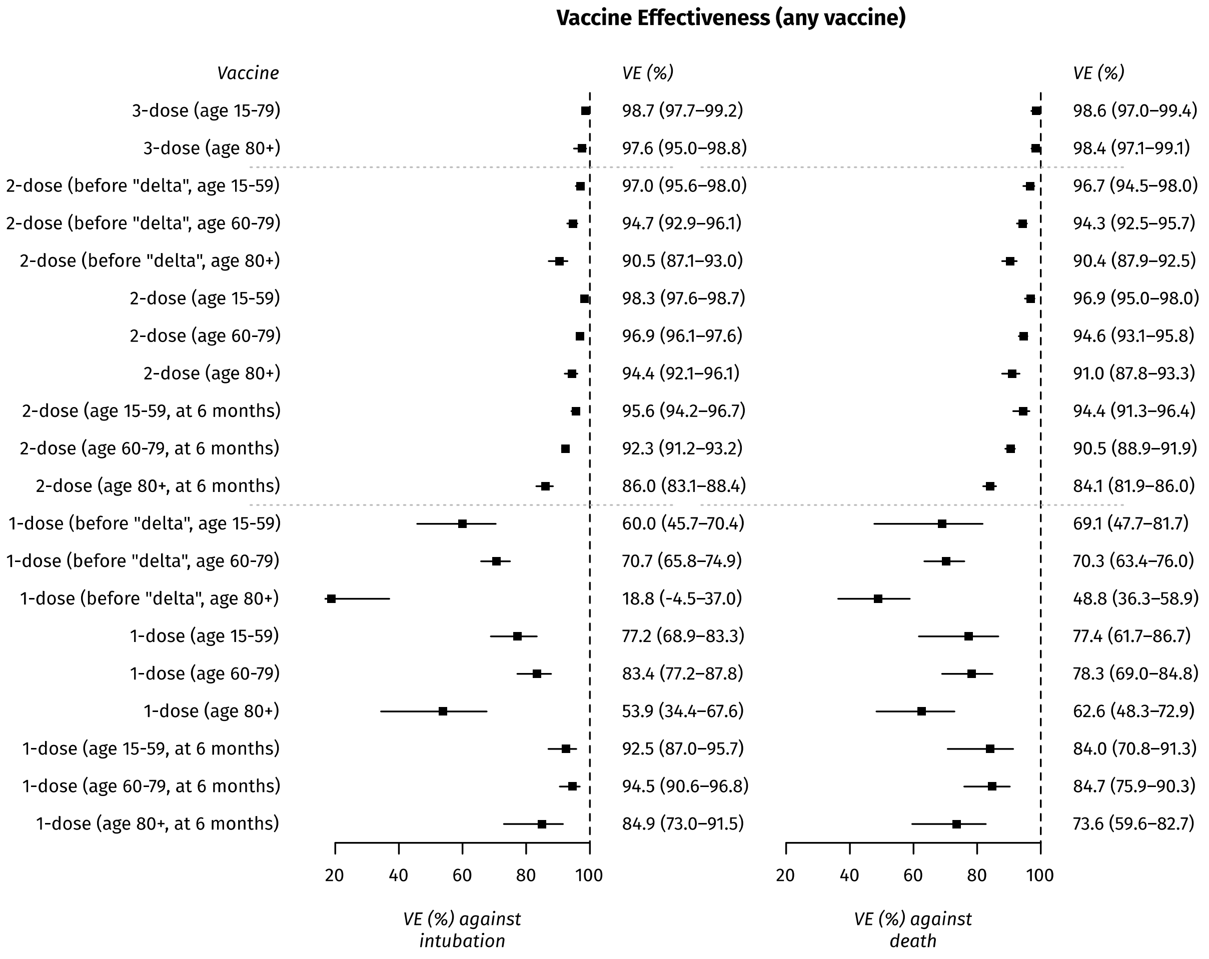


*(All results pertain to the "delta" variant, unless otherwise indicated.)*

**Supplementary Figure 2:** Comparative effectiveness of BNT162b2, mRNA-1273, ChAdOx1 nCoV-19 and Ad26.COV2.S vaccines against COVID-19 death and intubation, Greece, January-December 2021 (full results – model B, complete vaccinations)

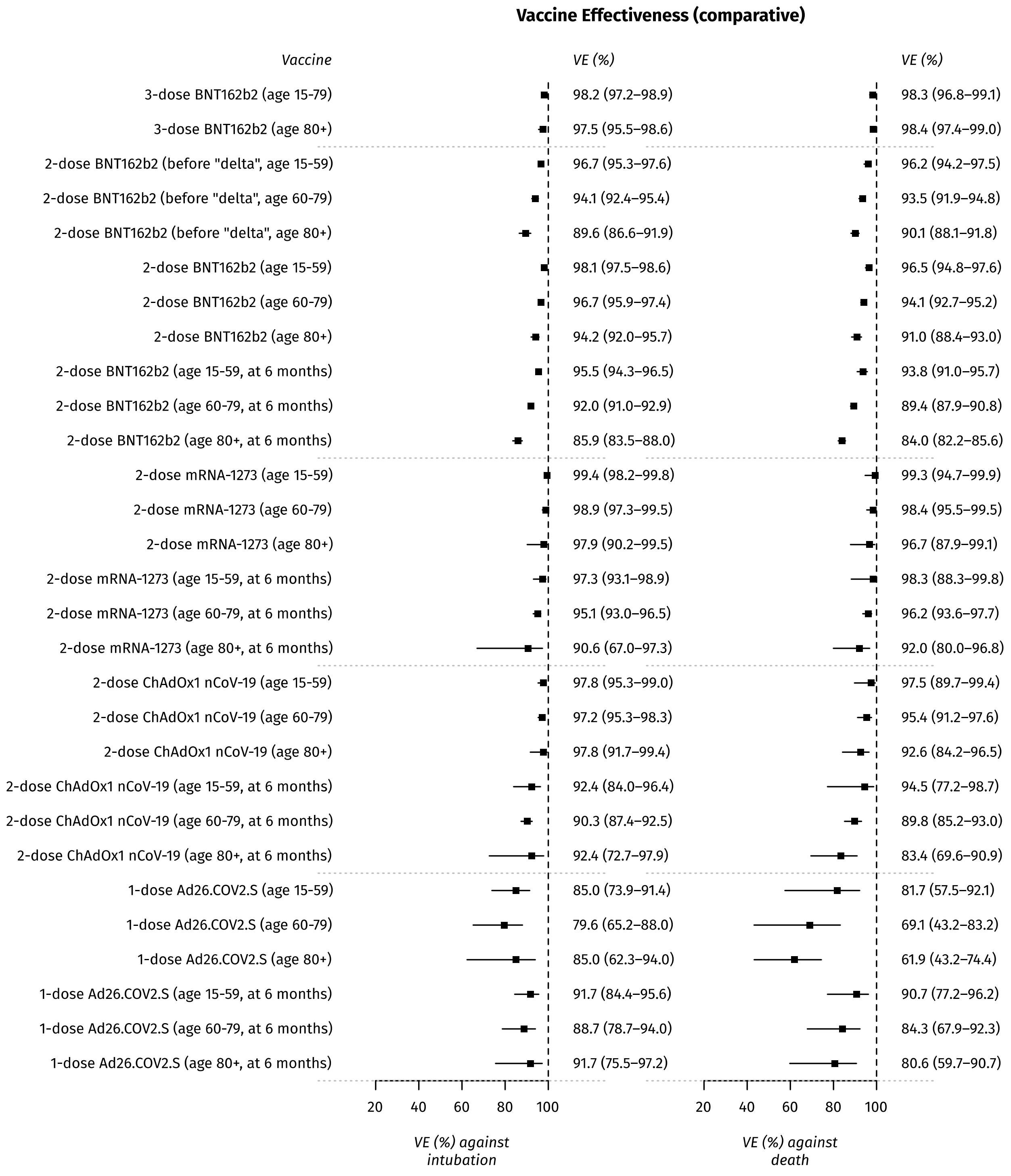


*(All results pertain to the "delta" variant, unless otherwise indicated.)*

**Supplementary Figure 3:** Comparative effectiveness of BNT162b2, mRNA-1273, ChAdOx1 nCoV-19 and Ad26.COV2.S vaccines against COVID-19 death and intubation, Greece, January-December 2021 (full results – model B, incomplete vaccinations)

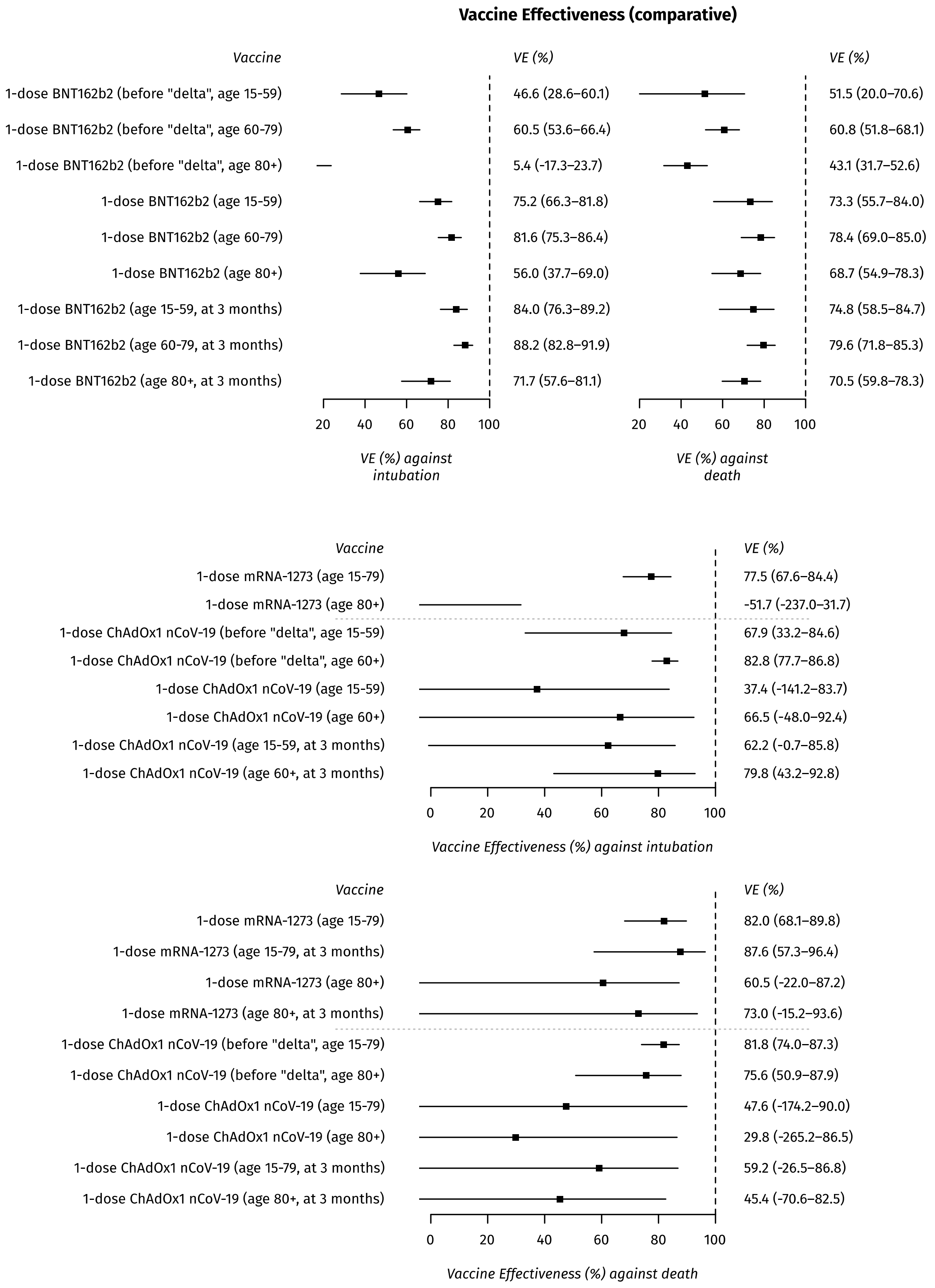


*(All results pertain to the "delta" variant, unless otherwise indicated.)*
